# Automated quantitative analysis of peri-articular bone microarchitecture in HR-pQCT knee images

**DOI:** 10.1101/2024.05.20.24307643

**Authors:** Nathan J. Neeteson, Sasha M. Hasick, Roberto Souza, Steven K. Boyd

**Affiliations:** Department of Biomedical Engineering, University of Calgary, Calgary, Canada; McCaig Institute for Bone and Joint Health, University of Calgary, Calgary, Canada; Hotchkiss Brain Institute, University of Calgary, Calgary, Canada; Department of Electrical and Software Engineering, University of Calgary, Calgary, Canada; Department of Radiology, University of Calgary, Calgary, Canada

## Abstract

There is growing interest in applying HR-pQCT to image the knee, particularly in the study of osteoarthritis, which necessitates the development and validation of novel image analysis workflows. In this work, we present and validate the first fully automated workflow for *in vivo* quantitative assessment of peri-articular bone density and microarchitecture in the human knee. Bone segmentation models were trained by transfer learning with a large dataset of radius and tibia images (N=2,598) and fine-tuned on a knee image dataset (N=131), atlas-based registration was used to identify medial and lateral contact surfaces, and morphological operations combined these intermediate outputs to generate peri-articular regions of interest (ROIs) for morphological analysis. Accuracy was assessed with an external validation dataset (N=131), where predicted and reference morphological parameters showed excellent correspondence (0.86≤R^2^ ≤0.99), with moderate bias present in predictions of subchondral bone plate density (-80 mg HA/cm^3^) and thickness (+0.15 mm). Precision was assessed with a triple-repeat measures dataset (N=29), where the short-term precision RMS%CV estimates ranged from 0.7% to 3.5% when rigid registration was used to synchronize ROI generation across images.

## Introduction

High-resolution peripheral quantitative computed tomography (HR-pQCT) is an *in vivo* medical imaging modality that produces density-calibrated images with a nominal isotropic resolution of 60.7 µm, allowing for direct and standardized measurement of bone density and microarchitecture^1^. HR-pQCT was originally intended for measuring bone at peripheral skeletal sites, primarily at the ultra-distal radius and tibia^2^. However, there is growing interest in utilizing this technology to measure bone quality in more proximal sites, such as the knee^3,4^ and the elbow^5^. Data processing and analysis procedures for HR-pQCT radius and tibia images are well-established^6^, but extending this technology for use at other sites requires the development and validation of new tools and protocols.

Our lab developed and validated a protocol for analyzing the peri-articular bone in the knee using HR-pQCT^7^ and has so far employed it in two cross-sectional studies^8,9^ and one longitudinal study^10^. Briefly, we summarize the analysis steps^7^: First, the image is segmented, using the standard semi-automated HR-pQCT segmentation procedure^11,12^ to separate the background, trabecular bone, and cortical bone (which we define to also include the subchondral bone plate, for simplicity). Next, the articular contact surfaces on the tibial plateaus and femoral condyles are manually identified. Finally, the articular contact surfaces are axially extruded into the bone to define four peri-articular regions of interest (ROIs) per compartment: subchondral bone plate and shallow (0 – 2.5 mm), middle (2.5 – 5 mm), and deep (5 – 7.5 mm) trabecular bone. These ROIs are shown in a volume rendering of the femur and tibia from a sample HR-pQCT image in Figure 1. In each subchondral bone plate ROI, the subchondral bone plate bone mineral density (Sc.BMD) and thickness (Sc.Th) are measured. In each trabecular ROI, the trabecular bone mineral density (Tb.BMD), thickness (Tb.Th), separation (Tb.Sp), and number (Tb.N) are measured, using standard methods^6^.

**Figure 1.**
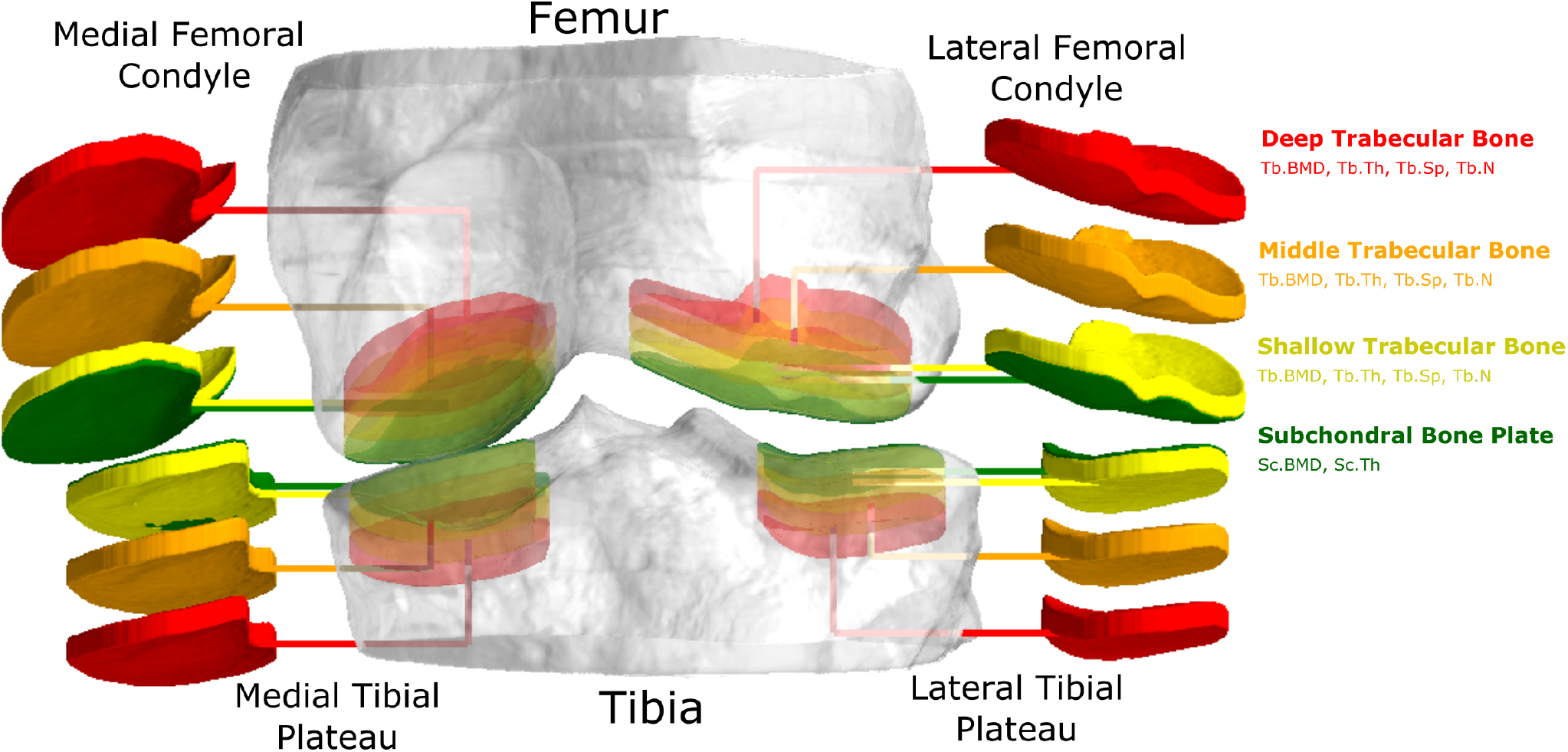
The peri-articular ROIs. The four compartments each have three trabecular ROIs (shallow - yellow, middle - orange, deep - red) and one subchondral bone plate ROI (green). The ROIs are labelled for the lateral femoral condyle, these labels also correspond to the same-coloured ROIs in each other compartment.

A major limitation of this protocol is that the semi-automated bone segmentation and the manual definition of articular contact surfaces incur a significant labour burden, not least due to the enormous size of a knee HR-pQCT image relative to the size of a more typical HR-pQCT image of the ultra-distal radius or tibia, hand, or foot. Furthermore, the subjectivity of these manual processes is a potential source of inter- and intra-operator errors and biases in study data^13^. A high labour burden limits the feasibility of scaling knee HR-pQCT studies to larger sample sizes, and inter-operator biases limit the ability to compare data between studies, scale studies to longer time periods, or conduct multi-center studies; all cases where data would be processed by multiple individuals and be subject to inter-operator biases.

Two well-established techniques for automating biomedical image segmentation are deep learning^14^ and atlas-based segmentation^15^. Fully convolutional neural networks, such as the UNet and its variants^16^, have been employed for segmentation to great success across many biomedical imaging modalities, including HR-pQCT^17,18^. In recent years, generative-adversarial^19^ and transformer-based^20^ architectures have also shown excellent performance for segmentation in both general and biomedical domains. Similarly, atlas-based segmentation is a well-established technique for biomedical image segmentation and has been applied successfully across several modalities and sub-domains^15^. While atlas-based segmentation is perhaps most well-known in the subdomain of neuroimaging, it also has a rich history of application in the subdomain of musculoskeletal imaging^21^.

The objective of this study is to develop and validate the first robust, fully automated protocol for peri-articular analysis of HR-pQCT knee images. We use deep learning for bone segmentation, atlas-based segmentation for articular contact surface labelling, and traditional morphological image processing operations to combine these segmentations and obtain the final peri-articular ROIs. We leverage several external validation datasets to evaluate the precision and accuracy of the developed protocol. This automated analysis protocol, which we intend to freely distribute, will accelerate HR-pQCT knee research by enabling studies to scale more efficiently and eliminating inter-operator biases in reported study data.

## Methods

### Datasets

Images used in this study were obtained using second-generation HR-pQCT (XtremeCT II, Scanco Medical AG, Brütisellen, Switzerland). All participants provided written informed consent before data collection. The Conjoint Health Research Ethics Board approved data collection at the University of Calgary (REB15-2108). There were three distinct sets of data used in this study: *training* data, external *validation* data, and *precision* data. Training data was used to train bone segmentation models and develop knee atlases. External validation data was used to assess the accuracy of the automated workflow, and precision data was used to quantify the precision of the automated workflow. All images in the external validation and precision datasets were scored for motion^22^, and images with a motion score of 4 or 5 (on a 1-to-5 scale) were excluded from the analysis.

### Training Data

There were two subsets of training data used in this study: the *distal leg/wrist* training data, and the *knee + background* training data. The *distal leg/wrist* training data contains pooled image data from two prior studies: a normative study (n=1,236)^23^ and a hip fracture study (n=108)^24^. These images were collected at standard scan sites for the distal radius and tibia, with 168 axial slices per image and a nominal isotropic resolution of 60.7 *µ*m. These images have reference segmentations labelling background, cortical bone, and trabecular bone, which were generated using the standard semi-automated protocol^6^. The full 3D images were preprocessed to create a training dataset consisting of 64x64x64 patch samples. Five patches were sampled from each full image with 50% of patches centered on cortical bone and 50% of patches randomly sampled from the full volume.

The *knee + background* training data contains image data from a previous longitudinal knee HR-pQCT study (n=19)^8^ investigating the effect of ACL injury on bone microarchitecture. In this study, each participant had bilateral HR-pQCT imaging of their knees at baseline (following ACL injury) and up to three follow-up visits. These images were collected following the protocol developed in our lab: the knee is placed at full extension in the scanner gantry and 1008 axial slices are collected, covering approximately 40 mm of the femur and 20 mm of the tibia, with a nominal isotropic voxel resolution of 60.7 *µ*m. Reference segmentations had been previously generated following the standard semi-automated protocol^11,12^ as well as articular contact surface masks and peri-articular ROIs generated^7^. The 3D images were preprocessed to create a training dataset consisting of 64x64x64 patch samples. From each articular contact surface in each image (medial femur, lateral femur, medial tibia, lateral tibia), 100 patches were extracted with 50% of the patches centred on the subchondral bone plate ROI and 50% of the patches randomly sampled from the vicinity of the trabecular peri-articular ROIs.

All knee images from prior studies had their background masked out as part of the data curation protocol that was followed at the time of those studies. To ensure the bone segmentation model could properly segment the background in new images, background patch samples were randomly sampled from 115 radius and 115 tibia images selected from the normative study dataset. From each of these images, five patches were sampled, centred on the background only. These background patches were pooled with the preprocessed knee image patches to create the *knee + background* training dataset.

### External Validation Data

The external test data contains pooled image data from three prior cross-sectional knee HR-pQCT studies: a cadaveric validation study (n=12)^25^ and two *in vivo* studies of bone microarchitecture in individuals who had undergone ACL reconstruction (n=58^9^, n=70^8^). These images were collected following the same protocol as the knee images in the training data, and have peri-articular ROIs generated following the same semi-automated protocols.

### Precision Data

A new repeat-measures dataset was collected to allow quantification of short-term precision of measurement workflows in knee HR-pQCT data. Adult participants were recruited using convenience sampling with a target sample size of 30^26^. Participants were screened for pregnancy at the time of recruitment by participant self-disclosure. HR-pQCT knee scans were performed on their non-dominant knee three times, with repositioning between scans, within a two week period. Knee HR-pQCT imaging followed the established protocol; however, to limit the total radiation dose, each scan contained only three stacks of 168 axial slices covering only the proximal tibia (excluding the distal femur).

### Bone Segmentation

#### Model Zoo and Training Procedure

Five distinct model architectures were considered for bone segmentation, all of which were 3D UNet variants or derivatives. Four models were imported from MONAI^27^: UNet^28^, UNet++^29^, UNETR^20^, and SegResNetVAE^30^. An additional fifth model, SeGAN^19^, was custom-implemented with PyTorch v1.12.1(+cu116)^31^. These specific models were selected due to their availability in an open-source package, their documented state-of-the-art segmentation performance, and because they each represent a distinct variation on the classic fully convolutional segmentation model architecture. All of the segmentation models take a 3D image with a single channel (rescaled densities) as input and produce a 3D image with three class channels (subchondral bone plate, trabecular bone, background) as output.

Training and validation were performed using five-fold cross-validation (CV)^32^. Models were trained using the PyTorch Lightning framework v1.9.4^33^ with a cross-entropy loss function, the AdamW optimizer^34^, and a batch size as large as would fit in memory for a given model using two NVIDIA A100 (2 x 80GB VRAM) GPUs, which varied based on model architecture and hyperparameters. In each fold, training proceeded for a minimum of 40 epochs with the following stopping conditions: four hours of wall clock time elapsed since the start of training, 1000 epochs, or no improvement in validation set subchondral bone plate Dice similarity coefficient (DSC)^35^ over the previous 40 epochs. The ranges of hyperparameters considered for each model architecture are documented in the supplemental information (Table S2).

#### From-scratch Learning

A hyperparameter grid search was performed with each of the five model architectures using five-fold CV with the *knee + background* training dataset. The best-performing model of each architecture was selected based on which had the highest mean validation set subchondral bone plate DSC. This resulted in five sets of five trained models: one model from each fold for each architecture. These were labelled the “from-scratch” trained models.

#### Transfer Learning

A hyperparameter grid search was performed with each of the five model architectures using five-fold CV with the *distal leg/wrist* training dataset. The best-performing model of each architecture was selected based on which had the highest mean validation set subchondral bone plate DSC (classifying radial and tibial cortical bone with the subchondral bone class). The models were reinitialized with the optimal hyperparameters and retrained using the full *distal leg/wrist* training dataset to convergence to be used as the pre-trained models. The pre-trained models were fine-tuned using five-fold CV with the *knee + background* training dataset. This resulted in five sets of five trained models, labelled the “transfer” trained models.

#### Ensemble Inference

Ensemble inference uses a set of models to produce a single prediction^36^. Each model performs patch-based inference on a full image using MONAI’s sliding window inferer^27^ with a patch width of 128 voxels, an overlap of 25%, and Gaussian blending. In our case, the five models are ensembled by summing their raw outputs element-wise before converting the outputs to voxel-wise class labels by finding the channel index with the largest predicted value at each voxel in the output. Ensemble inference is performed using a single NVIDIA V100 (16GB VRAM) rather than two A100s, as less memory is required for inference than for training.

### Segmentation Post-processing

Figure 2 shows a schematic of the segmentation post-processing workflow, including specific details of the morphological filters applied in each step. In step 1, the outputs from a set of trained models are ensembled to create the raw segmentation, labelling voxels as either subchondral bone plate, trabecular bone, or background. In step 2, the two bone labels (subchondral and trabecular) are combined to create a single bone mask. In step 3, morphological filters fill any gaps in the bone mask. In step 4, morphological filters keep only the largest connected component of the bone mask, which eliminates both small, “disconnected islands” resulting from segmentation errors and removes any secondary bones (e.g. in a proximal tibia image, the proximal fibula and distal femur are considered secondary). In step 5, the minimum subchondral bone plate is extracted from the filtered bone mask by eroding by 4 voxels and subtracting the eroded mask from the original mask. The result is combined with the raw subchondral bone plate mask. In step 6, a morphological filter removes all but the largest connected component from the subchondral bone mask and in step 7, the trabecular mask is created by subtracting this subchondral mask from the filtered bone mask. In steps 8 and 9, the trabecular bone mask is morphologically filtered to fill in any interior gaps and subtracted from the subchondral bone plate mask to ensure no overlap. Finally, in step 10, the trabecular and subchondral bone plate masks are combined into the final post-processed segmentation. All morphological and Boolean operations are performed using scikit-image v0.19.3^37^ and NumPy v1.21.6^38^.

**Figure 2.**
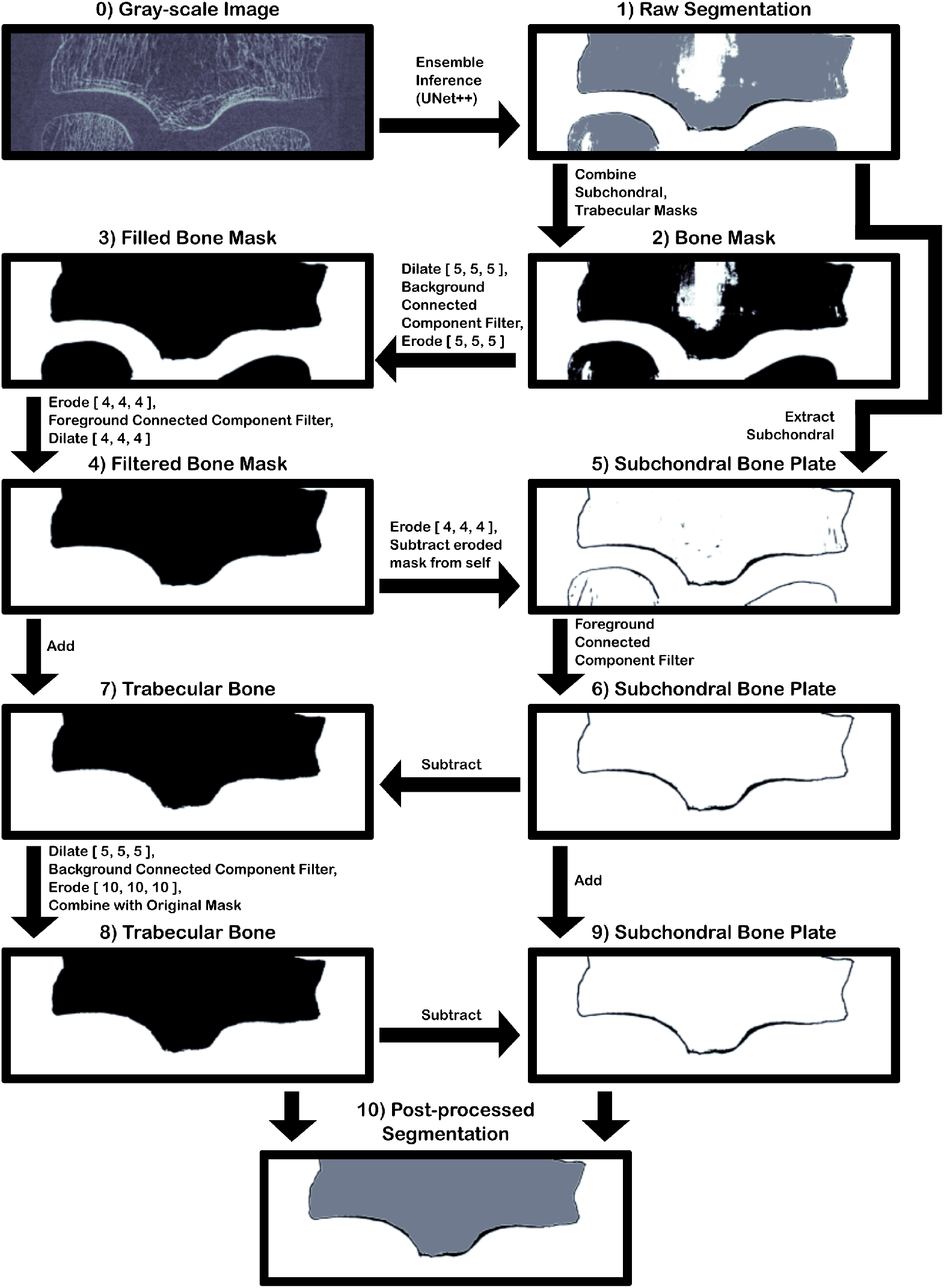
Segmentation post-processing workflow. Steps are shown as black arrows while intermediate outputs are visualized as 2D slices of the full 3D volume between steps. The workflow is demonstrated on a tibia with low bone density to demonstrate how the post-processing steps correct possible errors in the initial inferred segmentation.

### Atlas-Based Segmentation of Contact Surfaces

Articular contact surfaces on the tibial plateaus and femoral condyles are labelled on new images using atlas-based segmentation, with a separate atlas for each of the tibia and femur. The average atlases were created using the baseline knee images from the *knee + background* training dataset, which have manually generated masks defining the medial and lateral articular contact surfaces on the tibia and femur. First, images of left knees were reflected across the sagittal plane and combined with images of right knees so that all the images could be used to generate a single atlas (one for the tibia and one for the femur). An initial image is selected randomly and all other images are affinely registered to the initial image and averaged together to create the average atlas. Each of the original images is deformably registered to the average atlas and the contact surface masks from each image are transformed into the atlas space and combined to create the average atlas mask using the STAPLE algorithm^39^ with a binarization threshold of 0.5. The contact surfaces can be segmented in a new image by deformably registering the tibia and femur to their corresponding atlas and transforming the atlas mask to the new image. If the new image is from a left knee, the image is reflected sagitally before registration and the transformed masks are reflected sagitally back to the original orientation as the final step.

The optimal parameters for atlas generation and deformable registration were determined using a grid search (Table S1) and leave-one-out cross-validation, where there are as many folds as samples in the dataset. In each fold, one image was held out for validation while the rest were used to generate an atlas. The atlas was used to predict the contact surface mask on the validation image and the DSC of the reference and predicted mask were computed, which were averaged across folds. The set of parameters that produced the highest cross-validation DSC was considered optimal and used to generate a final atlas for the tibia and femur. The ranges of parameters considered in the atlas generation grid search are documented in the supplemental information, and the optimal procedure is reported in the Results section.

### Combined Workflow

The full combined workflow is depicted in Figure 3 for both cross-sectional and longitudinal analysis modes. The steps are described in full detail in the proceeding sections.

**Figure 3.**
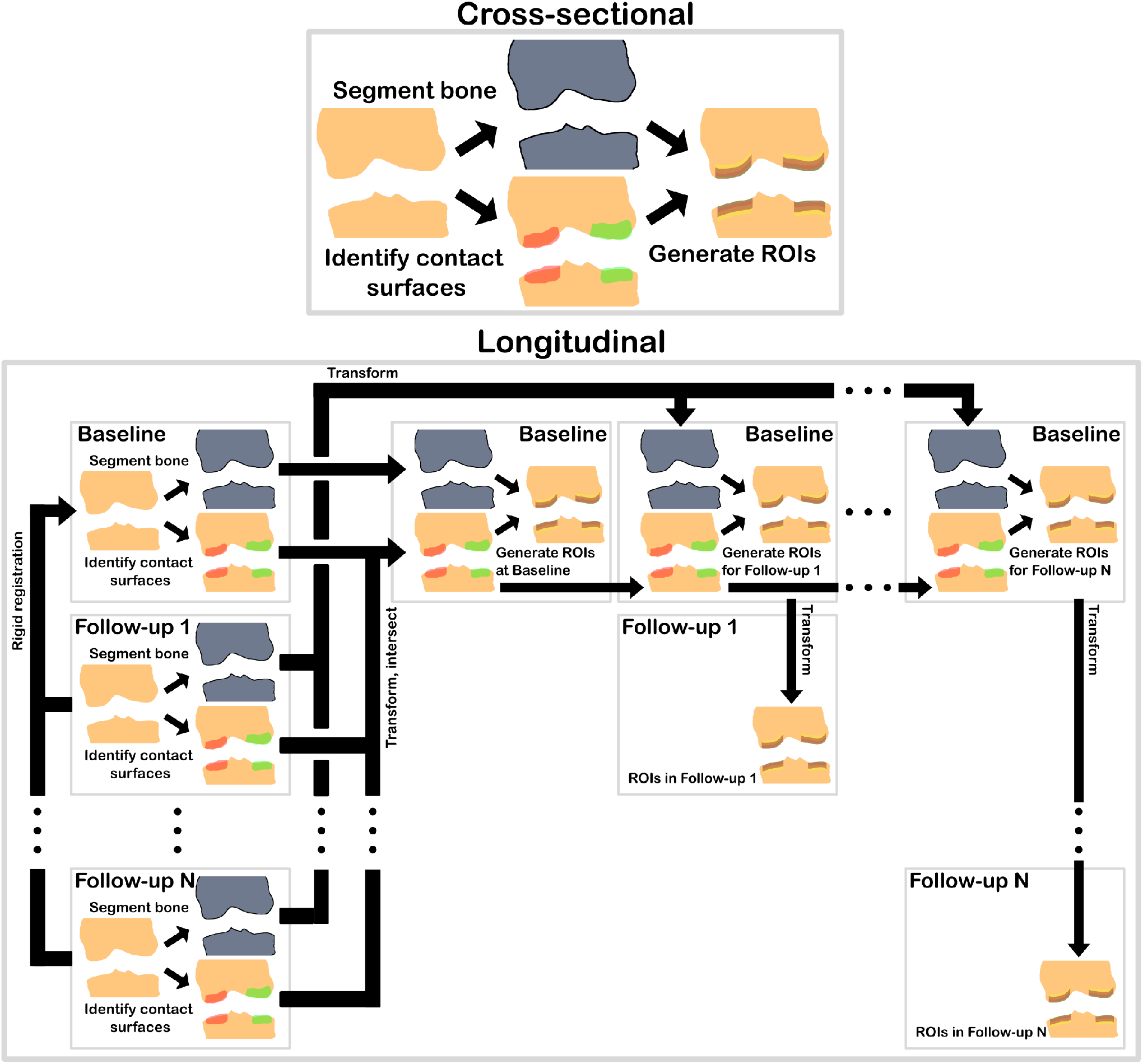
The combined workflow to generate peri-articular ROIs from a knee HR-pQCT image. With cross-sectional data, ROIs are generated for individual images. With longitudinal data, peri-articular ROIs are generated for each image in a time series using rigid registration. The bone segmentations and contact surface masks are transformed to the baseline frame to synchronize both the spatial location of the contact surfaces and the direction of the extrusions that generate the trabecular ROIs.

#### Segmentations

First, a full knee image is manually separated into two sub-volumes, using a custom script integrated into the manufacturer-provided GUI program for image analysis (*µ*CT_Evaluation, v6.6, Scanco Medical AG, Brütisellen, Switzerland). The two sub-volumes contain the distal femur and proximal tibia, which are processed independently. Ensemble inference and post-processing are applied to obtain a bone compartment segmentation. Next, the medial and lateral contact surface masks are obtained by atlas-based segmentation using the corresponding atlas.

#### Longitudinal Registration

If the image data is longitudinal (i.e., repeated-measures), each follow-up image is rigidly registered to the baseline image before generating the peri-articular ROIs. For rigid registration, fixed and moving images are first downsampled by a factor of 4 and Gaussian smoothed with a standard deviation of 1 voxel. The rigid registration uses a Powell optimizer with a maximum of 100 line iterations, a step length of 1, a step tolerance of 10^*−*6^, a value tolerance of 10^*−*6^, a geometry-based initialization, linear interpolation, a correlation similarity metric, a regular sampling strategy, and a sampling rate of 30%. Multi-scale registration is performed with downscaling factors of 16, 8, 4, 2, 1 and corresponding Gaussian smoothing standard deviations of 8, 4, 2, 1, 0.1 voxels. Longitudinal registration is performed using SimpleITK v2.2.0^40^. After registration, the follow-up bone segmentation and atlas-segmented contact surface mask are transformed to the baseline image space. The baseline and transformed follow-up contact surface masks are intersected to create the longitudinal contact surface masks in the baseline frame. This is combined with the bone segmentations, in the baseline space, to generate peri-articular ROIs for baseline and follow-up images. Follow-up peri-articular ROIs are transformed back to their respective follow-up space(s). If the image data is cross-sectional and only one image is available from each knee, the longitudinal registration step is skipped, and peri-articular ROIs are created using the atlas-segmented contact surface mask.

#### Peri-articular ROI Generation

A Gaussian filter (standard deviation: 1 voxel) is applied to smooth the medial and lateral contact surface masks. The smoothed contact surface masks are dilated axially out from the bone by 20 voxels (proximally for the tibia and distally for the femur) and intersected with the subchondral bone plate mask from the bone segmentation, and a connected components filter is applied to keep only the largest component. The resulting mask is dilated axially out from the bone by another 5 voxels and intersected once more with the subchondral bone plate mask from the bone segmentation. This creates the first two peri-articular ROIs: the medial and lateral subchondral bone plate. As in prior studies, the articular contact surfaces are extruded axially 7.5 mm into the bone (proximally for the femur and distally for the tibia) to create three trabecular peri-articular ROIs at three ranges of depth from the articular contact surface: shallow (0.0 – 2.5 mm), middle (2.5 mm – 5.0 mm), and deep (5.0 – 7.5 mm). This results in a total of eight ROIs in each of the femur and tibia.

#### Micro-architectural Analysis

Subchondral bone mineral density (Sc.BMD) and thickness (Sc.Th) are computed for the subchondral bone plate ROIs, while trabecular bone mineral density (Tb.BMD), thickness (Tb.Th), separation (Tb.Sp), and number (Tb.N) are computed for the shallow, middle, and deep peri-articular ROIs. All micro-architectural parameters are computed using standard manufacturer-implemented algorithms (IPL)^6^.

### Evaluation and Metrics

First, the combined workflow was applied to all images in the held-out test dataset in cross-sectional mode. The predicted and reference ROIs were used to perform micro-architectural analysis. The paired reference and predicted outputs were compared using both direct linear correlation analysis and the difference and mean values in Bland-Altman plots.

Next, the combined workflow was applied to all images in the precision dataset in both cross-sectional and longitudinal mode (using the first scan as the baseline and repeat scans as follow-ups). Generated ROIs were used to perform micro-architectural analysis and the cross-sectional and longitudinal results were used to compute the root-mean-square percentage coefficient of variation (RMS%CV) and least significant change (LSC)^26^ for all parameters at all ROIs in the tibia. The normality of the distribution of individual-level standard deviations was assessed with D’Agostino and Pearson’s method^41^ and the significance of differences in precision between the cross-sectional and longitudinal modes was tested using independent Wilcoxon signed-rank tests^42^, with the significance threshold adjusted for multiple testing using the Benjamini, Krieger, and Yekutieli two-step false discovery rate correction procedure (34 tests)^43^. The base significance threshold for all statistical tests was 0.05. Visualizations were created using PyVista v0.38.6^44^, Matplotlib v3.5.3^45^, pandas v1.3.5^46^, seaborn v0.12.2^47^, and Inkscape.

## Code Availability

The code used in this project is distributed across three GitHub repositories: utility code for training segmentation models in the PyTorch Lightning framework with HR-pQCT images (https://github.com/Bonelab/bonelab-pytorch-lightning), code for training, inference, and atlas-based segmentation (https://github.com/Bonelab/hrpqct-knee-segmentation), and code for organizing and analyzing the precision dataset (https://github.com/Bonelab/triknee).

## Results

### Datasets

The *distal leg/wrist* training dataset contained 2,598 total images (1,257 radii, 1,341 tibiae) from 1,276 participants (457 men, 819 women). The *knee + background* dataset contained 131 total images (each containing the femur and tibia) from 31 participants (10 men, 21 women). The external validation dataset contained 128 images from 64 participants (21 men and 43 women) and the precision dataset contained 87 images from 29 participants (10 men, 19 women).

### Segmentation Models

Figure 4 shows the mean cross-validation DSC for each class on the *knee + background* dataset for the best-performing version of each of the five architectures considered and with each training approach. All the best models achieve DSC≥0.94 at segmenting background and trabecular bone, except for the from-scratch trained SegResNetVAE. As expected, segmentation of subchondral bone plate is the most challenging aspect of the bone segmentation task. Most models achieved only 0.84≤ DSC≤0.87, the from-scratch trained SegResNetVAE achieved DSC = 0.66 (worse result), and the transfer trained SegResNetVAE and UNet++ achieved DSC = 0.89 (best results). Finally, the SegResNetVAE, UNet, UNet++, and UNETR show better performance across all classes when transfer trained than when trained from-scratch, while the reverse is true for the SeGAN. The transfer trained UNet++ was selected over the SegResNetVAE as the final model for bone segmentation as it had a lower standard deviation in subchondral bone plate DSC during cross-validation (UNet++: 0.889 ± 0.006, SegResNetVAE: 0.889 ± 0.014).

**Figure 4.**
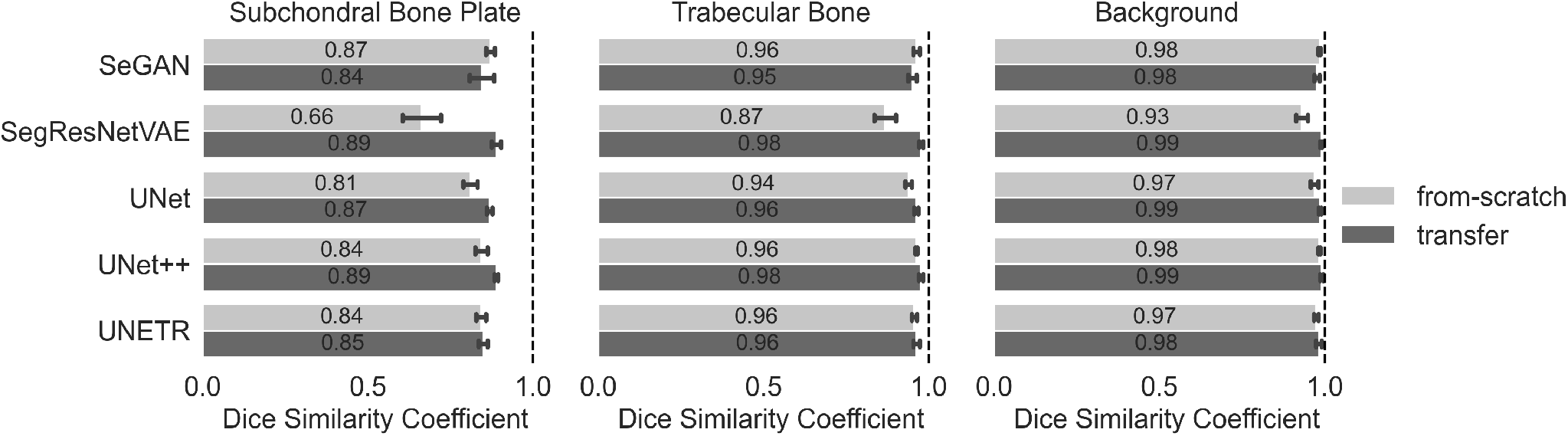
Cross-validation results for the best-performing model of each architecture trained with each method. The bars show the mean DSC averaged across folds, while the error bars show the standard deviation of the DSC. The dashed line indicates DSC = 1.0.

### Atlas-Based Segmentation

The best-performing atlas-based segmentation method was the diffeomorphic demons algorithm with a displacement smoothing standard deviation of 2 and an update smoothing standard deviation of 2. Images are downsampled by a factor of 8 and smoothed with a Gaussian filter with a standard deviation of 0.5 prior to registration, and multiscale registration is performed with down-sampling factors of 16, 8, 4, 2 and corresponding Gaussian smoothing standard deviations of 8, 4, 2, 1. The average DSC values across the leave-one-out cross-validation for this optimal configuration were 0.81 and 0.83 for the medial and lateral tibia plateaus, respectively, and 0.83 and 0.84 for the medial and lateral femoral condyles, respectively.

### Peri-articular microarchitectural parameters

Running times for the various steps in the workflow are reported in Table S3. Ensemble inference is performed on an NVIDIA V100 (16GB), and all other steps are performed on the CPU.

#### Accuracy

Figure 5 shows Bland-Altman and correlation plots comparing the results of morphological analysis on the external validation dataset with the reference and predicted peri-articular ROIs. For all morphometric parameters in all trabecular ROIs, the limits of agreement contain zero mean bias error, and the predicted and reference values are highly correlated (R^2^≥0.97). In the tibial subchondral bone plate ROIs, the limits of agreement overlap with zero, indicating no bias for both Sc.BMD and Sc.Th. In the femoral subchondral bone plate ROIs, the limits of agreement do not overlap with zero and indicate over-prediction of Sc.Th (+0.15 mm, compared to an approximate range of 0.35 to 0.85 mm) and under-prediction of Sc.BMD (-80 mg HA/cm^3^, compared to an approximate range of 500 to 800 mg HA/cm^3^). Predicted and reference values are highly correlated for Sc.BMD in the femur (R^2^ = 0.95) and Sc.Th in the tibia (R^2^ = 0.94), while the correlations are weakest, though still quite strong, for Sc.BMD in the tibia (R^2^ = 0.86) and Sc.Th in the femur (R^2^ = 0.88).

**Figure 5.**
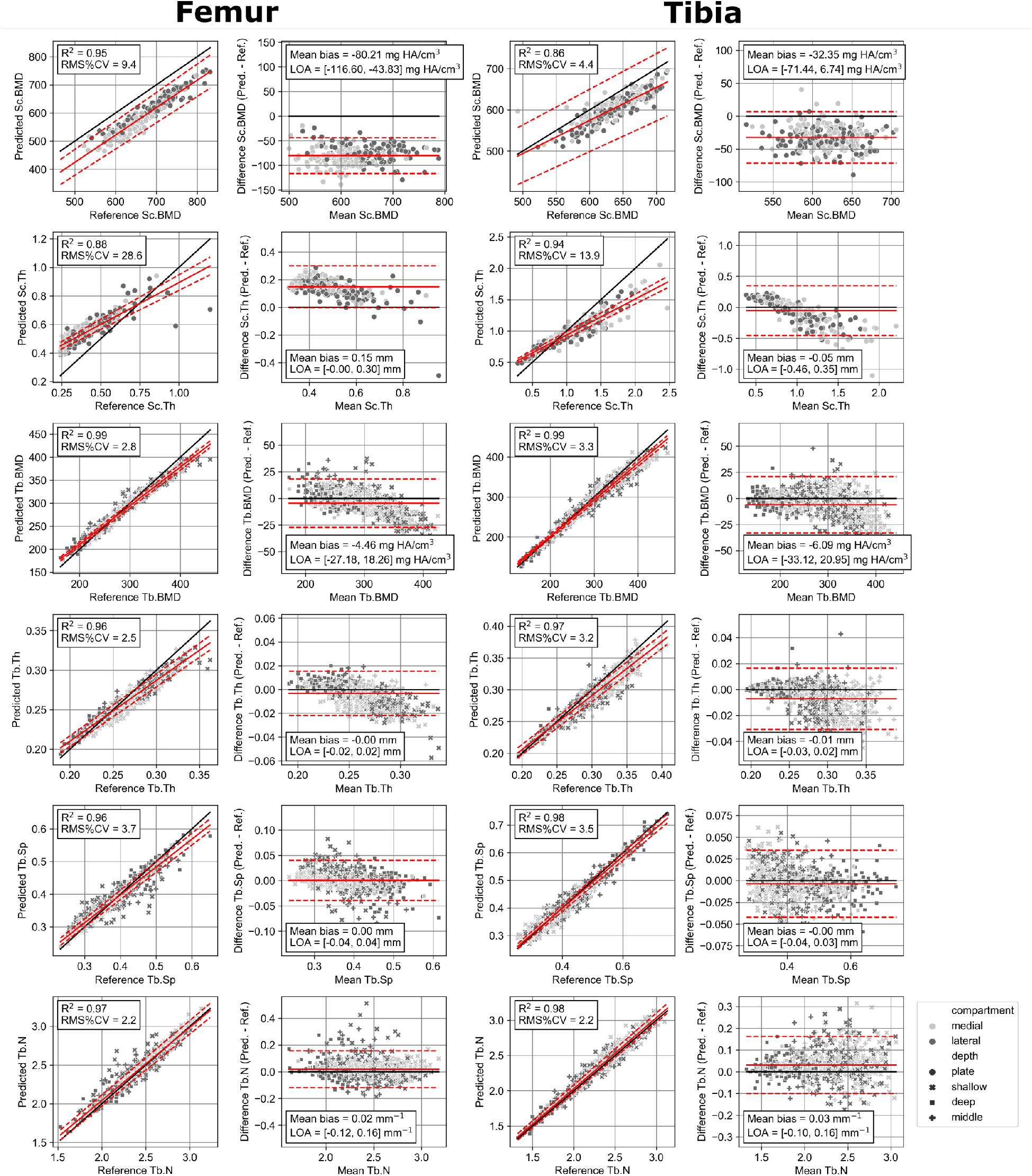
Linear correlation and Bland-Altman plots showing results on the external test datasets. On the linear correlation plots, the black line is y=x, the solid red line is a regression line, and the dashed red lines indicate the confidence interval for the regression. On the Bland-Altman plots, the black line is zero bias, the solid red line is the mean bias, and the dashed red lines are the limits of agreement.

Figure 6 shows sample visualizations of the compartments with the largest positive errors, the largest negative errors, and the median absolute disagreement in Sc.Th in the femur and tibia, with one row per image. There are two potential sources of disagreement: articular contact surface labelling, and subchondral bone plate segmentation. In each image, both types of error are present, with substantial disagreements at the medial and lateral edges of the image as well as at the endosteal boundary, which separates cortical bone, or subchondral bone plate, from trabecular bone. Notably, disagreements at the periosteal boundary are minimal, and correspondingly the differences at inter-ROI boundaries are also minimal. Finally, both the axial projections and sagittal slice overlays show good qualitative agreement between the predicted and reference ROIs, even for these four images with the largest errors in Sc.Th.

**Figure 6.**
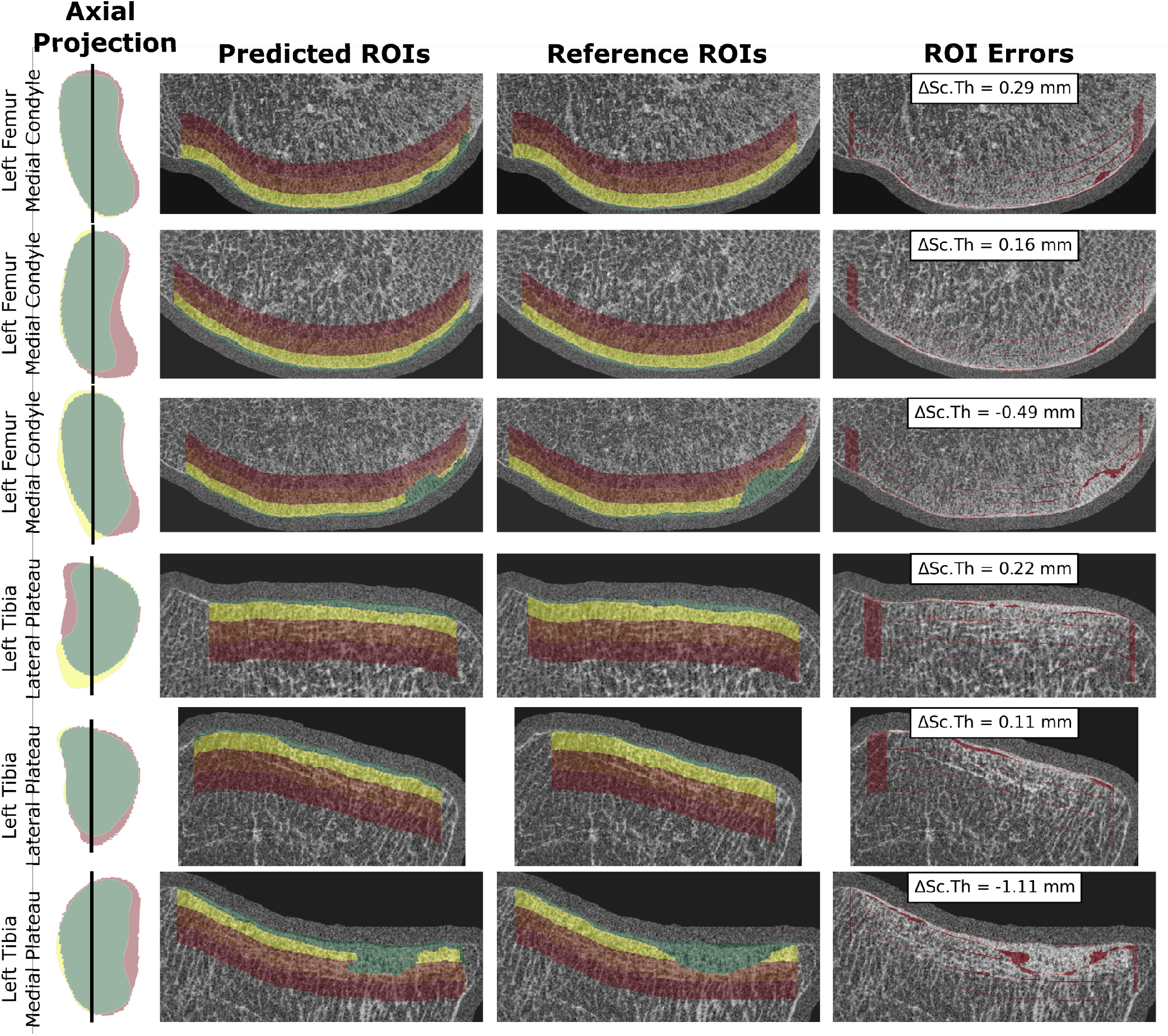
Sample visualizations of the compartments with the largest over-estimation of Sc.Th, median absolute disagreement in Sc.Th, and under-estimations of Sc.Th in the femur (top three rows, respectively) and tibia (bottom three rows). In the left-most column we show axial projections of the predicted and reference ROIs, combined into a single compartmental ROI, where green represents true positives, red represents false positives, and yellow represents false negatives. In the two center columns, the predicted (left) and reference (right) ROIs are overlaid on the gray-scale image of a central sagittal slice (the location of which is indicated by a black line in the left-most column). Here, green is the subchondral bone plate and yellow, orange, and red are the shallow, middle, and deep trabecular bone ROIs, respectively. ROI errors are shown in red in the right-most column and are defined as any voxel where the predicted and reference ROI labels disagree.

#### Precision

Table 1 contains RMS%CV and LSC values for all morphological parameters in all peri-articular ROIs in the tibia, computed in both cross-sectional and longitudinal analysis modes with the repeat-measure precision dataset. RMS%CV values are ≤3.5% for all parameter-ROI combinations, indicating excellent reproducibility of morphometric measurements across repeat imaging with repositioning. Precision is best for Tb.BMD (0.8% to 1.5%) and Tb.Th (0.7% to 1.3%), while Sc.Th shows the relative worst precision (2.2% to 3.5%). For all but two parameter-ROI combinations, the differences in precision between cross-sectional and longitudinal analysis modes are not statistically significant. The exceptions are Sc.Th in the medial tibial plateau, where the precision is significantly improved with the longitudinal mode (RMS%CV improves from 3.5% to 2.2%) and Tb.N across all trabecular ROIs, where precision is statistically significantly worse with the longitudinal mode, though the effect is quite small (RMS%CV worsens from 2.8% to 2.9%). For nearly all ROIs across all parameters except Tb.N, the trend is for precision to be improved when the analysis is performed in the longitudinal mode compared to the cross-sectional mode, while this trend is reversed for Tb.N.

**Table 1.** RMS%CV and LSC values for all microarchitectural parameters in all compartments of the tibia, compared between the cross-sectional and longitudinal modes. Differences in RMS%CV between modes were tested using Wilcoxon signed-rank tests.

| Parameter [units] | ROI | Cross-sectional |  | Longitudinal |  | Wilcoxon p Value |
| --- | --- | --- | --- | --- | --- | --- |
|  |  | RMS%CV | LSC | RMS%CV | LSC |  |
| **Sc.BMD**<br>[mg HA / cm<sup>3</sup>] | **All** | 2.2 | 37.3 | 2.2 | 36.6 | 0.980 |
|  | **Lateral** | 2.2 | 38.2 | 2.3 | 39.5 | 0.957 |
|  | **Medial** | 2.1 | 36.4 | 1.9 | 33.5 | 0.957 |
| **Sc.Th**<br>[mm] | **All** | 3.4 | 0.079 | 2.8 | 0.063 | 0.242 |
|  | **Lateral** | 3.4 | 0.066 | 3.3 | 0.066 | 0.980 |
|  | **Medial** | 3.5 | 0.091 | 2.2 | 0.059 | 0.035<sup>a</sup> |
| **Tb.BMD**<br>[mg HA / cm<sup>3</sup>] | **All** | 1.3 | 9.1 | 1.1 | 8.1 | 0.730 |
|  | **Shallow Lateral** | 1.5 | 12.4 | 1.2 | 10.9 | 0.803 |
|  | **Middle Lateral** | 1.4 | 8.8 | 1.3 | 8.7 | 0.980 |
|  | **Deep Lateral** | 1.3 | 5.8 | 1.2 | 5.4 | 0.961 |
|  | **Shallow Medial** | 0.9 | 9.6 | 0.8 | 8.4 | 0.980 |
|  | **Middle Medial** | 1.2 | 9.9 | 1.0 | 8.4 | 0.980 |
|  | **Deep Medial** | 1.2 | 6.6 | 1.0 | 5.4 | 0.250 |
| **Tb.Sp**<br>[mm] | **All** | 2.8 | 0.038 | 2.9 | 0.039 | 0.242 |
|  | **Shallow Lateral** | 2.6 | 0.032 | 2.7 | 0.030 | 0.458 |
|  | **Middle Lateral** | 3.1 | 0.045 | 3.2 | 0.044 | 0.659 |
|  | **Deep Lateral** | 2.7 | 0.043 | 2.7 | 0.044 | 0.980 |
|  | **Shallow Medial** | 2.6 | 0.028 | 2.6 | 0.027 | 0.957 |
|  | **Middle Medial** | 3.2 | 0.043 | 3.5 | 0.047 | 0.242 |
|  | **Deep Medial** | 2.3 | 0.036 | 2.4 | 0.037 | 0.980 |
| **Tb.Th**<br>[mm] | **All** | 1.0 | 0.0086 | 1.0 | 0.0085 | 0.242 |
|  | **Shallow Lateral** | 1.2 | 0.0099 | 1.2 | 0.0092 | 0.980 |
|  | **Middle Lateral** | 1.0 | 0.0072 | 0.9 | 0.0071 | 0.250 |
|  | **Deep Lateral** | 0.9 | 0.0059 | 0.9 | 0.0059 | 0.957 |
|  | **Shallow Medial** | 1.3 | 0.0123 | 1.3 | 0.0114 | 0.980 |
|  | **Middle Medial** | 1.0 | 0.0092 | 1.2 | 0.0106 | 0.250 |
|  | **Deep Medial** | 0.7 | 0.0048 | 0.7 | 0.0048 | 0.730 |
| **Tb.N**<br>[mm<sup>-1</sup>] | **All** | 2.8 | 0.15 | 2.9 | 0.16 | 0.035<sup>b</sup> |
|  | **Shallow Lateral** | 2.5 | 0.16 | 2.5 | 0.17 | 0.250 |
|  | **Middle Lateral** | 3.2 | 0.17 | 3.3 | 0.18 | 0.327 |
|  | **Deep Lateral** | 2.9 | 0.14 | 3.0 | 0.14 | 0.980 |
|  | **Shallow Medial** | 2.2 | 0.14 | 2.3 | 0.16 | 0.241 |
|  | **Middle Medial** | 3.2 | 0.18 | 3.4 | 0.19 | 0.358 |
|  | **Deep Medial** | 2.6 | 0.13 | 2.6 | 0.13 | 0.980 |

p values were corrected for multiple testing using the Benjamini, Krieger, and Yekutieli two-step false discovery rate correction procedure with a significance threshold of 0.05 and 34 tests.
<sup>a</sup> Precision is statistically significantly better with longitudinal method.
<sup>b</sup> Precision is statistically significantly better with cross-sectional method.

## Discussion

We have developed and validated a robust, automated protocol for peri-articular analysis of bone microarchitecture with HR-pQCT knee images. We found that of the segmentation models considered, the UNet++ trained by transfer learning achieved the most accurate segmentation of the subchondral bone plate in five-fold cross-validation, though many of the models performed nearly as well, while atlas-based registration by the diffeomorphic demons algorithm produced preliminary compartmental segmentations with DSCs of 0.81 to 0.84 in leave-one-out cross-validation. Using a held-out external validation test set, we found excellent accuracy and precision in standard bone morphometric analysis outputs when compared to the previous labour-intensive analysis workflow.

This is the first study to present an automated protocol for the analysis of peri-articular bone morphometry with *in vivo* HR-pQCT knee images and lays the foundation to leverage this technology more widely. While the application of HR-pQCT for studying bone quality in the human knee is relatively recent, there have been several studies demonstrating semi-automated protocols for both *in vivo* and *ex vivo* HR-pQCT knee images. These include the *in vivo* study of knee microarchitecture in OA patients and controls by dividing the tibial plateaus into four compartments (anterior, central, posterior, and medial or lateral)^3,4,48^, the *ex vivo* study of resected human femurs by manually creating volumes of interest, based on visual inspection^49^, and the *in vivo* protocol developed in our lab and described in the introduction^7^.

Comparing predicted to reference morphometric parameters in the test dataset (Figure 5), we observe strong correlation and high accuracy for all trabecular parameters (R^2^≥0.96) while for the subchondral bone plate parameters, the correlations are still strong but relatively weaker (0.86≤R^2^≤0.95). Sample visualizations (Figure 6) for the femur and tibia images show one of the biggest challenges is measuring the Sc.Th. This is not unexpected because the subchondral bone plate is extremely thin, which is evident both visually in Figure 6 and in the ranges of Sc.Th in Figure 5 (femur: ≤ 0.25 Sc.Th [mm] ≤1.2; tibia: 0.25 ≤Sc.Th [mm] ≤2.5). An extremely thin structure combined with endemic measurement artifacts (e.g. motion, partial volume effects, etc.) presents a difficult segmentation problem, and this is worsened by the possible presence of ambiguities in the definition of the boundary of the structure such as those caused by adjacent thickened trabeculae and porosities and/or breaks in the subchondral bone plate. The smaller the structure, the greater the marginal effect will be of incorrectly labelled voxels on density and thickness measurements. Compared to the subchondral bone plate, the bone as a whole and the articular contact surfaces are much easier to segment accurately (see Figure 4) All these effects combine to explain the greater accuracy in the trabecular parameters when compared to the subchondral bone plate parameters.

It is important to keep in mind that the correctness of the reference segmentations used to train and evaluate the model is subjective. These segmentations were generated using a semi-automated procedure with a human operator intervening to visually inspect and correct the periosteal and endosteal boundaries where required. Corrections were made using a manufacturer-provided tool (*µ*CT_Evaluation, Scanco Medical AG, Brütisellen, Switzerland) in which surface contours are inspected and adjusted laborously. A particular challenge is that in the knee the articular contact surfaces are predominantly oriented parallel to the axial plane and the proprietary software does not allow editing contours in other more convenient orientations. As such, it is difficult to ascertain which of the reference or predicted segmentations is objectively more correct when disagreements arise, or whether the idea of an objectively correct subchondral bone plate segmentation is even meaningful. In practical usage, it is most important that a measurement has a sufficient degree of self-consistency, or precision, such that differences can be reliably detected across groups or across time. When we apply our novel technique to a triple-repeat-measures dataset with N=29, we find excellent measures of reproducibility across all parameters and all compartments: 0.7 *<* RMS%CV [%] *<* 3.5, compared to 0.3 *<* RMS%CV [%] *<* 3.0 for similar parameters measured by a similar automated workflow at the distal radius and tibia^18^. Importantly, these measures of precision include not just the effects of the segmentation workflow but also effects caused by repositioning and rescanning. By comparison, the semi-automated procedure used to generate the reference segmentations was evaluated for inter- and intra-operator precision by comparing analysis results on a single set of images either performed by one operator twice (intra) or by two operators separately (inter), and this resulted in intra-operator precision values of 0.5 *<* RMS%CV [%] *<* 3.5 and inter-operator precision values of 0.5 *<* RMS%CV [%] *<* 7.9^8^. The fully automated workflow we present in this work has, by construction, zero inter- or intra-operator precision error.

Finally, while we did not find an across-the-board improvement of the precision of using rigid registration for repeat measures, we did observe a trend towards lower precision error, particularly in Sc.Th. One of the main benefits of rigid registration is to ensure that the projection of the ROIs from the articular contact surface into the bone occurs in the same direction in each image. In our precision data, repositioning was well aligned between scans, but in cases where there is a greater difference in the knee angle between measurements, it is presumed that registration would be an important tool to maximize precision^50^. In our precision dataset, all participants were healthy and had no difficulty in keeping their knee close to 0° of flexion consistently in each measurement. This is often not the case for injured participants with limited knee mobility.

There are limitations to this study to be acknowledged. All study participants were recruited locally in Calgary, AB, Canada and therefore the geographic and ethnographic heterogeneity of the study population is limited. It is not expected that this will limit its application, but that remains to be confirmed. All images were obtained using the same XtremeCTII scanner and all of the reference segmentations were generated by only two operators (one for radius and tibia images and one for knee images). Due to limitations of radiation dose, the knee image data for precision quantification contained only images of proximal tibia and excluded the distal femur. There was also a relative paucity of knee images with reference segmentations to be used for training and testing, since the application of HR-pQCT to the study of the knee is quite a new development compared to the well-established application to the distal radius and tibia.

There are elements of the workflow that could be improved by the incorporation of additional tools and/or techniques, or alternative measurement approaches. While the segmentation and ROI generation is fully automated, currently a user first spends approximately two minutes placing bounding boxes to crop out the distal femur and proximal tibia from a full knee image. This step saves memory and computation time in subsequent steps and is an opportunity to perform basic visual data checking and metadata entry before processing (e.g., to ensure the image has been reconstructed properly, to perform motion scoring, to make sure the image is from the correct side, etc.). However, it could potentially be automated to further streamline data processing. So-called ‘stack shift’ artifacts are a problem in any HR-pQCT measurement produced by performing multiple sequential measurements and “stacking” them together, and knee images are no exception. These artifacts can be particularly troublesome if the stack shift occurs at or near the articular contact surface, but could be ameliorated by incorporating a stack shift-correcting registration approach^51^. The thickness of the subchondral bone plate is approaching, or at, the resolution limits of HR-pQCT. An alternative to the standard measurement technique of segmentation and estimation of Sc.Th by Hildebrand’s method for calculating structure thickness^52^ could be to utilize the dual step function model-fitting approach originally proposed for measuring cortical thickness of the proximal femur in clinical CT^53^. Additionally, multi-task learning approaches have been shown in recent years to dramatically improve the performance of deep learning models^54^, particularly as a solution to data scarcity^55^. Multi-task learning could be applied to further improve subchondral bone plate segmentation by simultaneously training a model to predict additional holistic or voxel-wise characteristics of the input image, such as the motion score, the subchondral bone plate and trabecular thickness map, the marrow space thickness map, morphometric parameters, or even strength-based parameters derived from finite element modelling.

## Conclusions

We have presented and characterized a novel workflow for automating the peri-articular analysis of bone at the knee with *in vivo* HR-pQCT images. The algorithm runs fully automatically, requiring no human input after an initial cropping step, and can be operated in either a cross-sectional or longitudinal configuration as needed for a given study. We have demonstrated the accuracy of the workflow when compared to the best available reference data, and have evaluated the short-term precision and shown that it compares favorably to the precision of a similar workflow for obtaining similar measurements on radius and tibia HR-pQCT images. In its current form, it could be integrated directly and seamlessly into a standard HR-pQCT data analysis workflow, and the code, atlases, and trained models have been made available for anyone to use. Future work will focus on the practical application of the workflow on clinical data to investigate osteoarthritis etiology.

## Data Availability

The raw image data used in this study are not publicly available.

## Acknowledgments

The authors acknowledge Drs. Danielle Whittier and Andres Kroker for collecting and curating the training datasets and the support and contributions of the staff and students of the Bone Imaging Laboratory at the University of Calgary. This work was supported financially by a grant from the Canadian Institutes of Health Research (CIHR) [PJT 162189] and a Training Graduate PhD Salary Award from Arthritis Society Canada [TGP-21-0000000093].

## Author contributions statement

N.J.N. conceptualized and implemented the method, conducted the experiments, performed the statistical analysis, and wrote the manuscript. S.M.H. recruited participants and organized collection of the new *in vivo* data, and processed the new data.

R.S. provided input into method conceptualization and investigation. S.K.B. provided input into method conceptualization and investigation, and provided supervision and project administration. All authors reviewed the manuscript.

## Supplemental Information

**Table S1.**
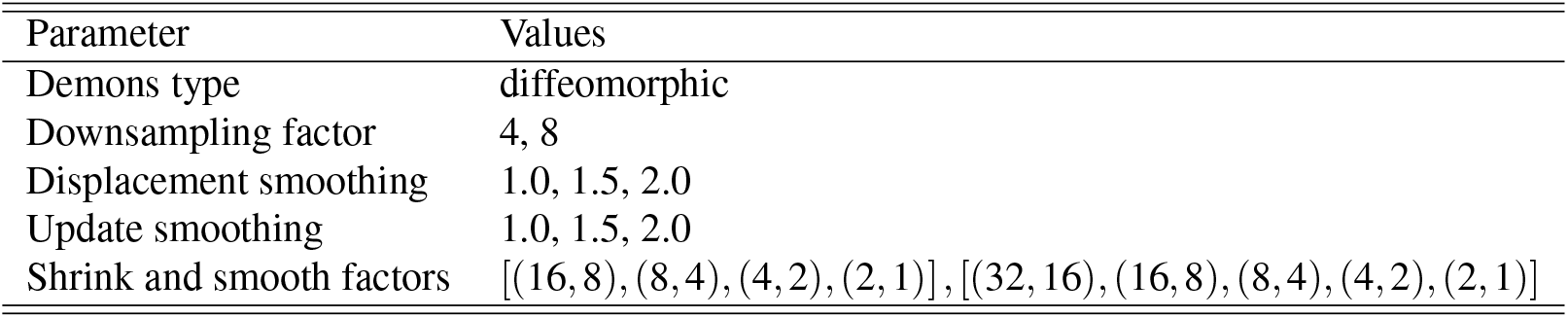
Parameter space for the deformable registration grid sweep. Only the diffeomorphic demons type was considered since it was important that the transformations be invertible. Shrink and smooth factors are reported as pairs of downsampling factors and Gaussian smoothing sigmas.

**Table S2.**
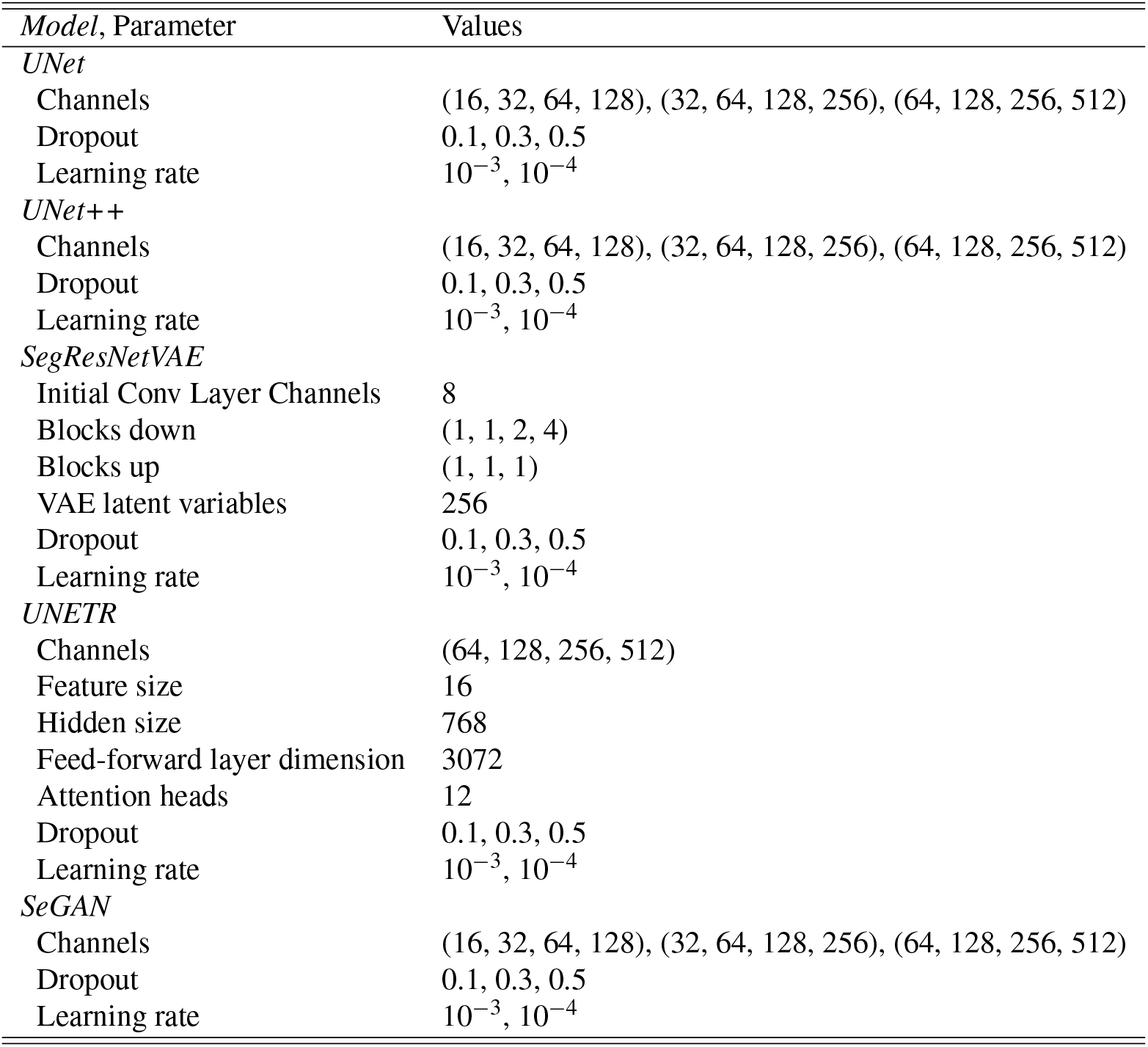
Parameter space for segmentation model selection when training from scratch on either training dataset. Parameter values grouped with round brackets indicate a single array of values.

**Table S3.**
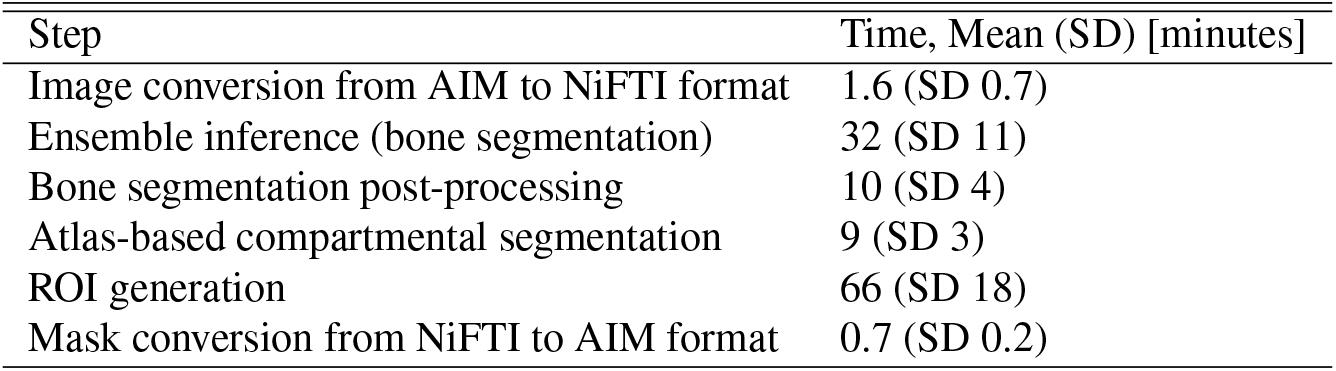
The running times for each step in the workflow were measured on the repeat-measures precision dataset and are reported as the average and standard deviation (SD) of running times for indnvidual images.

| Step | Time, Mean (SD) [minutes] |
| --- | --- |
| Image conversion from AIM to NiFTI format | 1.6 (SD 0.7) |
| Ensemble inference (bone segmentation) | 32 (SD 11) |
| Bone segmentation post-processing | 10 (SD 4) |
| Atlas-based compartmental segmentation | 9 (SD 3) |
| ROI generation | 66 (SD 18) |
| Mask conversion from NiFTI to AIM format | 0.7 (SD 0.2) |

